# A sense of being needed: an interpretative phenomenological analysis of hospital-based allied health professionals’ experiences during the COVID-19 pandemic

**DOI:** 10.1101/2020.11.18.20233908

**Authors:** Roel van Oorsouw, Anke Oerlemans, Emily Klooster, Manon van den Berg, Johanna Kalf, Hester Vermeulen, Maud Graff, Philip van der Wees, Niek Koenders

**Affiliations:** Radboud university medical center, Radboud Institute for Health Sciences, Department of Rehabilitation, Nijmegen, the Netherlands; Radboud university medical center, Radboud Institute for Health Sciences, IQ healthcare, Nijmegen, the Netherlands; Deventer Hospital, Department of Rehabilitation, Deventer, the Netherlands; Radboud university medical center, Department of Gastro-enterology and Hepatology-Dietetics and Intestinal Failure, Nijmegen, the Netherlands; Radboud university medical center, Donders Center for Neuroscience, Department of Rehabilitation, Nijmegen, The Netherlands; HAN University of Applied Sciences, Faculty of health and Social Studies, Nijmegen, The Netherlands

## Abstract

**Background:** The outbreak of COVID-19 has led to many hospitalizations in the Netherlands and worldwide. As a result, the work of hospital-based allied health professionals changed rapidly. In such a healthcare crisis, professionals may not be able to fulfill their own professional values regarding good care, resulting in ethical issues and moral distress. The aim of this study was to explore the lived experiences of allied health professionals working in hospitals during the COVID-19 pandemic, including the ethical issues and moral distress these professionals might have encountered.

**Methods and findings:** Lived experiences were collected in semi-structured interviews with 39 allied health professionals including dieticians, occupational therapists, physical therapists, and speech-language therapists, working in four different hospitals in the Netherlands. Interviews were held in June and July 2020, after waning of the first wave of COVID-19 spreading. Interpretative phenomenological analysis revealed four themes: ‘a disease with great impact’, ‘personal health and safety’, ‘staying human in chaotic times’ and ‘solidarity and changing roles’. Participant experiences show that the virus and COVID-19 measures had a significant impact on the in-hospital working environment due to the massive downscaling of regular care, infection prevention measures and unknown risks to allied health professionals’ personal health. At the same time, participants experienced a certain freedom, which made room for authentic motives, connection and solidarity. Participants felt welcomed and appreciated at the COVID-19 wards and intensive care units, and were proud that they were able to fulfill their roles. The themes and accompanying ethical issues reflect a wide range of situations that were morally complex and led to moral distress.

**Conclusions:** To diminish long-lasting negative impact of the COVID-19 pandemic and moral distress, employers should empathize with experiences of allied health professionals and create conditions for ethical reflection. Our data show that allied health professionals value professional autonomy. Creating room for professional autonomy makes them feel needed, connected and energized. However, the needs of allied health professionals may conflict with organizational rules and structures.

## Introduction

On the 11^th^ of March 2020, the World Health Organization (WHO) declared the outbreak of COVID-19 to be a pandemic. This outbreak, caused by the contagious novel Coronavirus (SARS-CoV-2), led to a large number of hospitalizations worldwide.[1] At the peak of hospitalizations in the Netherlands, in the beginning of April, over 1,300 patients with COVID-19 were admitted to intensive care units (ICU) and over 2,600 patients were admitted to hospital wards.[2] Hospitals in the Netherlands reached their capacity limits, especially in districts with a large number of COVID-19 cases. This situation caused severe challenges for hospital management and distress of health professionals.

During this first wave, patients hospitalized with suspected or confirmed COVID-19 were cared for in full isolation and separated from other patients, using “cohorting”. Cohorting allows dedicated staff to work in a ward only with COVID-19-positive tested patients, so they are unlikely to carry the virus from positive to negative patients. Press conferences from the Prime Minister and Minister of Health were held in order to announce measures in order to slow down spreading of the virus, and to “flatten the curve”.[3] Employees in vital professions were asked to keep working at their workplaces, whereas other citizens were advised to work from home or close their business. Schools and childcare were rapidly closed, causing children to be at home. Visiting policies in hospitals were adjusted and, in some cases, fully restricted. At the same time there were shortages of personal protection equipment (PPE).

Hospital-based allied health professionals (AHPs), including dieticians, occupational therapists, physical therapists, and speech-language therapists, were tasked with a variety of roles in the care of patients with COVID-19, in a working environment very different from normal. They had to make decisions with limited knowledge of optimal precaution and treatment strategies. Due to the crisis, the work process was often task-based instead of human-based and holistic. Although the professional roles were mostly well delineated, AHPs could face roles for which they were not prepared.

As earlier outbreaks have shown, decision-making in a time of emergency is associated with ethical issues.[4] Ethical issues occur when values and norms conflict, or when they no longer seem applicable. These conflicts may occur within a person, between different persons, or there may be disagreement with the rules or structures that guide his or her professional activities.[5] In these situations, it is not possible to avoid acting, therefore decisions have to be made.[6] Making difficult decisions in troubling situations such may result in moral distress.[7] Moral distress is defined as *“one or more negative self-directed emotions or attitudes that arise in response to one’s perceived involvement in a situation that one perceives to be morally undesirable”*.[8] Moral distress among healthcare professionals can cause professional dissatisfaction, burnout, and shortages in medical staff.[7] Nurses and physicians regularly face life-threatening and morally sensitive situations which explains why moral distress is extensively studied among these professions.[9] However, moral distress can be experienced by healthcare professionals across disciplines.[10]

During the COVID-19 pandemic, AHPs supported nurses and physicians, often beyond the scope of their normal professional roles, meaning that they might have faced moral distress as well. To our knowledge, the experiences of AHPs working in hospitals during a pandemic have not been studied yet. Insight into their experiences could help to empathize with their unusual working situation and identify requirements for healthy deployment of AHPs during this pandemic or a possible future one. The aim of this study was to explore the lived experiences of AHPs working in hospitals during the COVID-19 pandemic, including the ethical issues and moral distress these professionals might have encountered.

## Methods

### Qualitative approach and research paradigm

We performed an interpretative phenomenological study to explain lived experiences of AHPs in four hospitals in the Netherlands. Phenomenology, founded by Edmund Husserl, is a philosophical approach to study experiences.[11] It focuses on the appearance of the world to the individual, through *“going back to the things themselves”*. We collected first-person accounts of experiences with semi-structured interviews and analyzed the interview data with interpretative phenomenological analysis.[12] Reporting of this study followed the Standards for Reporting Qualitative Research.[13]

### Researcher characteristics and reflexivity

The interviews were performed by three researchers (RvO, NK and EK), all hospital-based physical therapists and involved in healthcare for patients hospitalized with COVID-19. The interviewers were all educated and experienced in performing qualitative research. Prior to the interviews, the interviewers bracketed their own experiences, opinions and prejudices on working as an AHP in the hospital during the COVID-19 pandemic.[14] They did so by writing a reflection report. Before start of the data analysis, the two data analysts (RvO and NK) bracketed their experiences, opinions and prejudices once more in a reflection report. The other researchers have a background as ethicist (AO), dietician (MvdB), speech and language therapist (HK), nurse (HV), occupational therapist (MG), and physical therapist (PvdW).

### Sampling strategy

Through purposeful sampling, AHPs were selected for participation in this study. Optimal variation was sought in gender, age, allied health profession, years of work experience, and type of hospital. Potential participants were eligible for study inclusion if they were employed in a hospital as a dietician, occupational therapist, physical therapist, or speech-language therapist and if they participated in healthcare for patients hospitalized due to COVID-19 infection. Potential participants were excluded from the study if the professional had no access to digital communication equipment or if the professional suffered from psychological complaints requiring professional support.

Team managers of allied healthcare departments in four hospitals in the Netherlands, two academic and two general hospitals, were asked to recommend potential participants. These professionals received an email from the author (NK) to provide study information and to invite them for participation. Professionals not responding to the invitation received one reminder email. Professionals willing to participate signed their informed consent digitally using Castor electronic data capture.

### Data collection

First-person accounts of experiences were collected to study the lived experiences of AHPs, setting aside assumptions and prejudices from participants and interviewers about common sense and science. Participants were asked to share their thoughts and experiences in one in-depth semi-structured interview of maximum 60 minutes. The interviews were structured by an interview guide covering the following topics: working in the hospital, working with patients with COVID-19, working at the cohort ward, working at the ICU; screening, diagnostics, treatment, ending treatment and referral of patients with COVID-19; personal protection equipment and infection precautions; ethical dilemmas, moral stress, and personal health. The interviewers stimulated the participants to describe the experiences as they had lived them and to avoid causal explanations, generalizations, or abstract interpretations.[15] All interviews were held though video calling to avoid the risk of viral spreading. The interviews were audio-recorded.

### Data analysis

All interviews were transcribed verbatim by a third party. Data were analyzed following interpretative phenomenological analysis, a thoroughly described method situating participants in their specific context, exploring their personal perspectives.[12] This method consists of six steps: step one to five were performed independently by the data analysts (RvO and NK), step six was performed by all authors. *1) Reading and re-reading a transcript*. For this step the transcript of one interview was read and the recording was listened simultaneously. *2) Initial noting*. In this step the interview was coded line-by-line, looking specifically for experiential claims, concerns and understandings of the case’s lived experiences. *3) Developing emergent themes*. The initial codes from step two were turned into themes through interpreting by the data analysts. The analysts looked for the essences of the things said. *4) Searching for connections across emergent themes*. The data analysts fit together the themes and selected the themes relevant to the research question. Connections between themes were noted in one case description per case. *5) Moving to the next case*. Step one to four were repeated for each case. *6) Looking for patterns across cases*. Patterns across cases were searched in two consensus meetings using the transcripts (step 1) and case descriptions (step 4). The two analysts presented and explained their case descriptions. All authors looked for similarities and differences between the two case descriptions. After three unique cases, they hypothesized patterns across cases by looking for connections, potential themes, certain orders, and key emergent themes. Finally, the first author (RvO) went through the individual case descriptions once more, to check whether they were adequately reflected by the established themes.

### Techniques to enhance trustworthiness

The transcripts were aggregated and analyzed using ATLAS.ti software (ATLAS.ti version 8.4, Scientific Software Development GmbH, Berlin) supporting open and transparent data analysis. All data and steps of data analysis can be traced back in the audit trail. Furthermore, credibility and trustworthiness of interviews were enhanced by the timing of the data collection. There was maximum three months between the lived experiences in the work situation and the interview. The collected data were analyzed by two different researchers independently to create maximum thoroughness and to correct for individual blind spots. By using a heterogeneous sample of four different types of AHPs, we created an interprofessional understanding of lived experiences. Moreover, the consensus meetings improved transparency and thoroughness of the data analysis. All quotes of participants were translated by a third-party native English speaker.

### Ethical considerations

As described in the introduction, caring in times of a pandemic can be intense and distressing. Therefore, the interviews concerning this work period were anticipated to be potentially sensitive, emotional or confronting. Participants were informed about the sensitivity of topics in the research information letter. At the start of the interview it was emphasized that the interview could be paused or stopped at any moment without giving any reason. Moreover, individual support was secured by all team managers. The Radboudumc ethical committee (dossier number 2020-6520) judged that this study does not fall under the scope of the Dutch Medical Research Involving Human Subjects Act (WMO). General principles from the Declaration of Helsinki and Good Clinical Practice were followed.[16,17]

### Context

The interviews were performed in June and July 2020. In these months, the numbers of hospitalizations concerning COVID-19 infected persons in the Netherlands were low, see Figure 1. The lockdown measures in the Netherlands were less restrictive starting May 2020. In June and July 2020, the basic rules still applied: stay home when being sick, work from home by default, wash your hands more often than usual and keep 1,5 meters distance. Public life had somewhat normalized, with students returning to schools, wearing facemasks in public transport and cafés and restaurants being open to a maximum of 30 persons.

**Figure 1.**
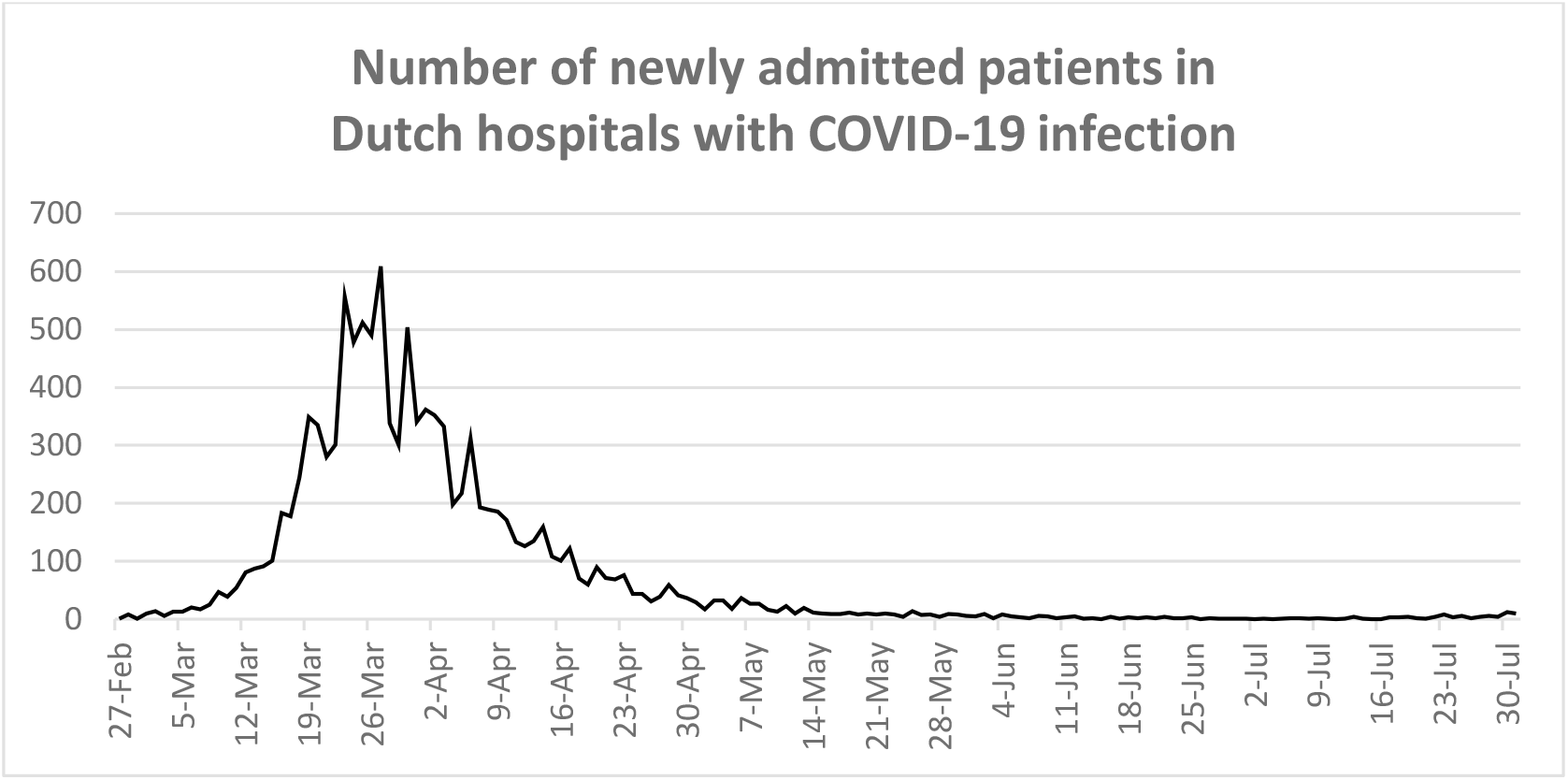
Number of newly admitted patients in Dutch hospitals with COVID-19 infection.

## Results

### Participants

Forty-two potential participants were contacted, three of whom did not respond to the study invitation. In total 39 AHPs enrolled in the study and completed the interview. The sample consisted of 9 dieticians, 7 occupational therapists, 13 physical therapists, and 10 speech therapists. Two of the 39 interviews were held through a phone call with audio recording without video connection, because of technical failure. Interviews had a mean duration of 48 minutes (standard deviation: 6 minutes). Participant characteristics are displayed in table 1.

**Table 1.** Participant characteristics.

| Participant characteristics (N=39) |  |
| --- | --- |
| Gender |  |
| Women | 29 (74%) |
| Men | 10 (26%) |
| Age (in years) |  |
| Mean (standard deviation) | 39 (11) |
| Range | 24 - 64 |
| Profession |  |
| Dietician | 9 (23%) |
| Occupational therapist | 7 (18%) |
| Physical therapist | 13 (33%) |
| Speech-language therapist | 10 (26%) |
| Work experience in the hospital (in years) |  |
| Mean (standard deviation) | 15 (11) |
| Range | 1 - 42 |
| Working in type of hospital |  |
| University hospital | 22 (56%) |
| General hospital | 17 (44%) |

### Theme descriptions

Four themes emerged from the data analysis: ‘a disease with great impact’, ‘personal health and safety’, ‘staying human in chaotic times’ and ‘solidarity and changing professional roles’. Thorough descriptions of the themes are provided below in order to elucidate their meaning. Participant quotes are used to exemplify particulars of the phenomenon. They are numbered in correspondence to the numbers in the open access database.[18] The theme descriptions contain ethical issues as experienced by the participants. Themes, ethical issues and accompanying values at stake are displayed in table 2.

**Table 2.** Themes, ethical issues and accompanying values at stake.

| Theme | Ethical questions | Values at stake |
| --- | --- | --- |
| A disease with great impact | How ought I treat patients without sufficient knowledge of the disease?<br>How can I avoid over-exerting patients? | Beneficence, non-maleficence, professionalism |
| Personal health and safety | Is it safe to work in the hospital?<br>Am I putting my loved ones at risk? | Patient health, personal health, health of family, responsibility, duty |
| Staying human in chaotic times | How do I keep a healthy work/life balance?<br>How do I stay human in these conditions?<br>How can I treat patients in a humane way? | Personal health, duty, humanity, dignity |
| Solidarity and changing professional roles | What role should I take on?<br>How can I support my colleagues?<br>Am I a general healthcare professional or a specialized allied health professional? | Responsibility, solidarity, competence |

### A disease with great impact

One of the first encounters of Dutch AHPs with COVID-19 was media footage of Italian ICUs with many patients mechanically ventilated in prone position. At the end of February 2020, the first case of COVID-19 in the Netherlands was confirmed. Participants pointed out that they knew many very sick patients could come their way and impact their work heavily, which made them apprehensive. In some cases, AHPs were trained and briefed to be able to help nurses. Non-acute care was scaled down and AHPs working in outpatient clinics had to call their patients to cancel appointments. Some participants experienced some very quiet shifts, in which they were waiting for patients to arrive. They sensed a quiet before the storm and a looming threat.

> “That was surreal. I came in, and it was actually very quiet from eight to ten and then between ten and twelve we suddenly had emergency admissions and scheduled admissions and that’s when you saw the nurses panic, because the nursing team had to be completely reorganized as they suddenly had six patients instead of three.” (participant 10320)

When the number of hospitalized patients increased, participants reported that things changed very quickly. Hospital wards were restructured in order to provide cohort treatment and nursing staff was shuffled around. When hospitals reached their patient ward limits, patients were transferred to other hospitals. Participants felt like they had ended up in a bad movie. There was a sense of disbelief and being taken by surprise.

> “During one of my shifts at the ICU a helicopter landed on the front lawn three times to take patients from the ICU to another hospital because we were full. It was also quite intense to see a long line of ambulances loading and unloading people.” (participant 36412)

The AHPs talked about a crisis mode, and among nurses and physicians, a survival mode. There was uncertainty and questions such as: What is happening? Will we be able to cope with all these patients? Participants explain that they felt anxious, especially those who witnessed severe illness due to COVID-19 among loved ones, or AHPs living in areas with a large number of patients with COVID-19. The streets were empty, because of the government measures, which made going to their work weird and surreal. Hospitalized patients were very sick, requiring high doses of oxygen, many patients were mechanically ventilated in prone position and many patients died. The participants were overwhelmed by the severity of the illness. Some patients had great impact on the wellbeing of participants, especially when patients were of similar age as themselves.

> “One of the most intense moments [I experienced] took place on that first Friday when a man my own age was being intubated. I was born in the year ‘79 so when I heard him come in and I heard the staff say that there would soon be a man admitted that was born in ‘79, I thought, oof, he is my age. […] In the end he did make it after 3 days. He was also off the ventilator after 3 days. I checked up on him for a while. Some people get under your skin a little.” (participant 10320)

The AHPs saw patients being lonely, and felt the urge to help and comfort them. This feeling was amplified by the fact that family and close relatives of patients were often forbidden from visiting as a result of infection precaution measures. AHPs saw patients in fear and panic and witnessed sad moments, which made them feel powerless. Concerning the treatment of patients with COVID-19, there was uncertainty about what to do. AHPs had little information about the disease. Participants did not dare to rely on their clinical experience because the disease showed an unpredictable course, very different from normal. They faced ethical issues: How can I treat patients without sufficient knowledge of the disease and its unpredictable course? How can I avoid over-exerting patients? AHPs tried to solve these ethical issues by being more careful, and by including more checks and balances. Some participants felt they were failing the patients, because they did not have the expert knowledge needed to effectively treat patients. The participants wondered if patients would ever recover, and what would be left of their quality of life.

> “Usually, if you look [at a patient] objectively, you would say that as he swallows so well, we can stop the treatment, but at the same time I think, yesterday he could not do it yet, why is that? So, I will make sure to check [on the patient] tomorrow and if he still does well, then I will stop my treatment. That sudden change in patients is something I do not recognize, and I think most of us do not, that is really something that we think is typical of COVID-19, that made it more difficult.” (participant 37470)
>
> “What really got to me is seeing patients panic, but not really being able to reassure those patients, to say don’t worry, it will all work out, we know what we are doing. When it comes to other patients in the hospital you often know the course [of illness] so you can also assume a more reassuring role. So I thought that was, well, I found it quite strenuous mentally.” (participant 38684)

The participants experienced a sharp contrast between being inside and outside of the hospital. From the outside, the situation seemed eerie due to the stories being told in the media. Being inside the hospital did put things into perspective according to AHPs. There was a feeling of dread, a feeling of pressure, but at the same an excitement before and while entering the wards. Participants typically mentioned they were curious and wondering: What is happening behind those closed doors? This made it even more rousing to go through the doors or sluices.

> “At one point it was like that for a few weeks and then I thought well, I would actually like to be more involved because I am getting afraid here at home, because I am not there (in the hospital). Because at home it all seems very spooky, but when you are there it is all very different.” (participant 84710)
>
> “I was a bit apprehensive because well, measures were taken, security staff was sitting by the door, people were scanned and those, well, other routes that you had to walk, certain departments that you were not allowed to enter, you do become a little curious, like what is happening over there?” (participant 48827)

The participants explained that they stepped in a different world when they entered the cohort wards or ICUs. The PPE measures impacted the look and feel of the wards, it created an unpleasant personal distance between patients and professionals and caused them to lose their ‘human touch’. In a short time, people were used to the situation inside. It was relatively quiet, in contrast to the chaotic context pictured in the media. The participants said they saw “with their own eyes that patients were still humans, irrespective of their condition”. Despite the isolation precautions, it was the scenery and work with which they were familiar. The AHPs noticed a difference between colleagues that had seen the wards from the inside and those who had not. The anxiety and feeling of threat were lower among those who had been on the inside according to the participants. They wished that the other colleagues would be allowed to see the patients and wards with their own eyes, because it would put things in perspective.

> “Actually, in the beginning I often thought of the images of Italy, with all that plastic and I don’t know, it being completely hectic, but when I actually got to the ICU it was quite tranquil, and there were patients in the rooms, but the doors were closed and there was no chaos at the time.” (participant 11036)
>
> “Of course, you have a kind of specter imagining people vomiting everywhere and having diarrhea and well, I don’t know really, but that was actually, at least, in my experience it was not too bad.” (participant 69932)
>
> “Once you are passed the sluice room, and you arrive at the department itself, well, that felt strange, but the kind of distress or madness that you see on television and in the media, was not present at all.” (participant 33394)

Some participants suddenly had to work from home; a change that was very abrupt and felt strange. The participants expressed feelings of emptiness, resignation, and guilt towards colleagues working in the hospital. Participants point out that it felt safe to be able to work from home, however, on the other hand it was disappointing that they could not take part in the clinical work. The possibilities to work remotely were seen as positive and offered good solutions. However, participants typically missed their connection with patients, certainly when they had to work with “patients from paper”.

> “It felt like you were about to be fired. Not that I was afraid of my dismissal, but it felt very weird. Because you sort of say goodbye to your colleagues and you do not know when you will see each other again. So, we ended up calling each other every day. We did plan a sort of meeting at the end of each working day and eventually it turned out that this was not necessary, instead we often connected by app and by mail. But it felt very strange.” (participant 84710)
>
> “You are a health care worker so you want to do your best, on the other hand, it is also a virus, that we can all, that we are all susceptible to and well, there are very strict measures in place and you need to stay at home and work from home as much as possible. Then perhaps being at home and not necessarily having to visit the hospital, gives you and your family a safe feeling.” (participant 46670)

### Personal health and safety

During the first COVID-19 wave in the Netherlands there was little information about the seriousness of the disease. There were stories that about 25% of the hospitalized patients would die. There were cases of healthcare professionals who got sick and were admitted to their own ICU. Furthermore, there was uncertainty about the way the virus spread. Participants point out that they were very aware of the danger of the virus, and that they took action to avoid becoming infected. They felt a great responsibility for the health and wellbeing of their loved ones. Participants faced the question: I want to help in the hospital but is it safe? If I do, will I put my loved ones in danger? This made participants meticulous regarding their hygiene. Suits were changed more often, hands were washed more often, keyboards were cleaned more regularly, and they hugged their children only after first taking a shower. Some participants chose to be in very strict self-isolation to solve this issue and stayed available for work in the hospital. Participants that had to work in the hospital while they had expressed concerns to their supervisors about being infected with the virus, felt guilt towards patients and colleagues who they might have exposed, and made them feel angry and frustrated.

> “That realistic fear that makes you think in the beginning, maybe I will get it, or maybe I will infect other patients, yes, I did have that.” (participant 30812)
>
> “Both my father and my mother are still alive. They are 86 and 87. Yes, I take care of them a lot more these days because they are not allowed to, and cannot, go to the shops themselves. So, I am very involved with my parents. I was involved already, but now even more so. And yes, I have been saying all this time that if they get sick and they die of COVID-19 then there is nothing we can do, because they are old and have little resistance. But I don’t want them to get it from me.” (participant 58414)
>
> “My wife is a little older than I am and she has some lung problems, so she might be susceptible, and I didn’t want to infect her. I know that at my age I am not a big risk to my kids, but the situation I am in means that I can catch it more easily because I am exposed all day, if I take proper hygiene measures, this also applies to my grandchildren and my parents. So yes, I consciously said that I am going to go into quarantine and stay 1.5 meters away from others at home and that is quite easy to do, but you have to be aware of it and we were [.…] I didn’t want to be the one to spread it, to infect vulnerable people and loved ones so that was my main motive.” (participant 51388)

In addition, close relatives of participants also raised concerns about the safety of their work. They asked whether participants formed a risk because they were in contact with infected persons and asked if the participants did not feel scared to take on face-to face contact with patients with COVID-19. Overall, participants felt safe using the PPE on the cohort wards and ICUs. Some stated that they were aware of the relatively luxurious position compared to the dearth of PPE in the primary care setting.

> “At one point I really felt much safer here in the hospital than in the supermarket. I was often asked if I was not afraid, and about wearing a suit, and how it must be quite stressful, but to be honest, I would rather be in a suit than in the supermarket.” (participant 37470)

Some situations caused participants to worry that they had been exposed to the virus, in particular when they witnessed disconnection of mechanical ventilation, when they questioned whether they had pressed the nose pads of the mask sufficiently, or when their skin was not fully covered by the PPE. One participant stated that she wore extra high socks to avoid skin contact. Some participants knew themselves to be hypochondriac, and recognized anxiety about becoming infected as a continuing threat. Many participants opted for a healthy lifestyle in order to be more resilient to the virus and to stay available for work in the hospital.

> “Then I agreed with my boyfriend not to drink alcohol anymore because we felt like we wanted to stay healthy, we both work with COVID-19 patients, so we stopped [drinking]. And we started exercising, three times a week, together with our colleagues. That worked out very well, because it stopped us from only talking about COVID-19 at work, and changed the subject to sports. Never in my life have I exercised so much, ate that healthy, and abstained from any alcohol. […] so strangely enough I came out fitter than I went in. No COVID-19 pounds.” [participant laughing] (participant 30952)

The AHPs felt anger towards people not following the rules, or not keeping the required social distance. This anger was aimed at people outside the hospital, for instance in the supermarket, and also at colleagues in the hospital. Participants feeling vulnerable were extra keen regarding the measures and felt troubled when colleagues were not as compliant, however, they did not want to be the one to keep correcting them. Other participants feeling vulnerable due to previous illness considered it as a personal victory that they were able to keep working in times of COVID. This made professionals feel strong and energetic.

> “For me it really is like I told you, about having been ill, and at that time, it was questioned whether I could ever lead a normal life. And that took about two years. And then I slowly started living a normal life again. But I never dared to dream that I could do this. And now I’ve done it, so now I feel like I can take on the world.” (participant 13293)

### Staying human in chaotic times

In a short period of time adjustments were made in the hospitals including restructuring departments for cohort, replacing staff, and implementing new treatment guidelines and protocols. Participants felt a need for clarity and leadership. Some participants recognized aspects from previous epidemics, such as coping with patients with HIV. The restructuring came with a continuous stream of information. There were many newsletters, emails and webinars to update the healthcare professionals. Participants pointed out that frequent changes in policy made the situation chaotic and they spoke of an overkill of information through all different channels. It was difficult to distinguish between the main and the side issues. In addition, it took much time to distil what information was actually relevant for their work.

> “I was a bit apprehensive because you had to deal with a lot of new rules. On how to act, how to put on and take off the insulation materials correctly, whether I was allowed in or not?” (participant 38684)
>
> “You don’t dare not to read it because you might miss something you need to know. So, I always read everything but, in the end, I thought hmm, does this actually make a difference for me?” (participant 58414)

Rules and policies changed over and over again. The AHPs experienced this period as chaotic and hectic, they had to be alert all day. This also concerned PPE usage. Patients had to be cared for in full isolation and at the same time scarcity of PPE material resulted in orders to avoid patient contact unless strictly necessary. AHPs felt pressure to not use any materials if not very urgent. Working with the PPE was physically tough: participants mentioned they felt dehydrated, and experienced headaches in the evening. They had to be creative in unfamiliar circumstances, avoid going in and out of rooms and departments, find solutions in the use of materials, because the normal instruments were not available at cohort wards and ICUs, or could not be used due to limited options for cleaning. Moreover, they had to work with inexperienced teams and colleagues, because nurses were oftentimes relocated. With many things different, participants were happy to recognize their colleagues on the work floor. Working in this new situation and with this new disease, participants explain that they could no longer rely on their routine, which made their workdays intense and exhausting.

> “Usually the door is open so you walk in and start treating someone and you occasionally do consult with the nursing staff of course, but that happened more often now because the door is closed and you don’t really want to waste materials [PPE], so you ask in advance, who’s behind the door? How are they doing? What should we do? What do we want to do? We need to make choices. So, we actually added a step [to the process] in order to choose what is necessary and what is not, and where I should go” (participant 58247)
>
> “Everyone was a little lost, there was a group of regular ICU nurses of course, MC [Medium Care] nurses from wards other than the ICU, OR [operation room] staff who were working on the ICU, nurses from the day center and yes, they had to be trained. They didn’t know where anything was, they hardly knew anything about the way the ICU operates, so yes, you noticed that it was different than usual and the regular ICU staff were also allocated to various departments, so you came into contact with other people as well when generally you are in your own bubble so to speak, you know everyone, you are familiar with each other and that was different now.” (participant 36915)

The period was experienced as intense and stressful, but on the other hand also valuable. After work they could feel tired, cranky and hot-tempered. Some participants experienced difficulty sleeping. They took their work home, and the experiences had to be processed. Participants were confronted with the question: I want to fulfill my job in the best possible way, however, how do I keep a healthy work/life balance? It helped to share stories and feelings with loved ones. Others explained that they had to write things down, or take time to ponder in order to process things. After the COVID period they felt tired and needed a period to cool off. Participants experienced the workload in different ways. Especially the participants with small children explained that is was a very busy period due to the home situation with home schooling. Others stated that the period was quite relaxed, because a lot of meetings were cancelled and appointments with friends and family were cancelled.

> “I also quite liked the fact that a lot of meetings that you usually had to attend had been canceled. So, it certainly has advantages. So yes, I do hope we can hold onto some things from this period of time.” (participant 26976)
>
> “If we could fully return to work immediately, I would certainly like that. On the other hand, I also see the benefits of working more flexibly and being able to make your own choices.” (participant 98542)

The unfamiliar chaotic situations, with all professionals dressed in PPE, and many sick, often sedated, patients with COVID-19 made the work at the wards and the ICUs feel less humane than normal. In the crisis there was a high turnover and a lot of patients died. Therefore, some participants felt that they blocked their emotions. They experienced a robot mode and the work felt factory-like. This typically was the case for AHPs participating in teams that supported ICU nurses turning patients in prone position. Participants felt that they were treating bodies, or human-like dolls, rather than persons. Some situations were referred to as disgraceful. Patients were oftentimes naked, smelling, bloated and affected by severe decubitus, which AHPs had not experienced before. There was a lack of personal information in the rooms, which made it impossible to provide person-centered care. This felt wrong, and some participants felt guilt towards those patients. They wondered: how do I stay human in these conditions? And how can I treat patients in a humane way? They coped with this by talking to patients, and by showing dignity and respect, even when patients were deeply sedated and were not expected to hear anything of the things said.

> “What really, well, what really affected me was the one nurse who addressed people by name and told them [what we were doing] we will now do this or do that. I really liked that, so I picked it up and started doing it myself. I started addressing people too. Otherwise, it feels like a rag doll lying there.” (participant 58414)

The AHPs wanted to restore and promote the patients’ dignity as much as possible. Some participants explained that they work in healthcare because they want to care for people. They feel the need for human contact and empathized with the lack of contact patients received.

> “People also feel really lonely don’t they, and as I said, all doors are closed, you have to rely on that one moment when someone comes in, all suited up, well then, that real contact, that real human contact is not there really and it is that human interest that I think we need to make sure we hold on to.” (participant 66521)

The participants stressed the importance of a personal connection when motivating patients. Participants found creative ways to establish human contact, despite the PPE boundaries. They sought eye contact more consciously and frequently, or used their voice in particular ways. Some participants explained that they very deliberately showed their face to the patient, before entering the room.

### Solidarity and changing professional roles

The period of crisis was experienced as a time of solidarity. Participants typically felt an urge to support their colleagues, to relieve busy physicians and nurses and to help keep the hospital up and running. Participants speak of a caring reflex, a duty, a calling, a feeling of commitment, a need to stand by when needed, to be of service. They felt drawn to the hospital. They felt that this was why they had become a healthcare professional. Many hospital-based AHPs were deployed as general healthcare professionals. Some helped transporting patients through the hospital, others were stationed at the nursing ward to support nursing staff. They helped with washing, replenishing stocks, bringing medicine and so on.

> “An event like this also creates solidarity. You go for it together and that is a great feeling.” (participant 30556)
>
> “It really felt like you just had to be there for the department, you know, and offer help. Because you knew there was a lot of pressure on the nurses and on the, well, the hospital. And if you can support them as a dietitian by simply being more accessible and also being there for them in the weekend, so that it does not become an issue for them, then I think that will also make their work easier. […] They should be able to focus on the primary process as much as possible.” (participant 83109)
>
> “On my first day off, I went cycling. That is my hobby, so I went for a leisurely ride, reducing stress. And I as cycled towards [city name], I was inclined to ride to the hospital. At that moment you realise that you are constantly taking on the assisting role. It didn’t matter what you did, either. I even bathed a patient; those are nursing duties. The nurse asked me if I wanted to bathe the patient, and I said I never did that before but—by then I’d also seen a lot of feces, and at that point that doesn’t matter as long as you can help the nurses. They are very busy.” (participant 10320)

Some participants state they felt frustration when they were not allowed to help on the wards due to rules or management choices. When coming to and working on the wards or ICUs they felt welcomed and appreciated. There was a strong sense of team spirit among healthcare workers. The participants felt proud that they were able to fulfill their role.

> “You just do it. In my opinion the risk you took was small compared to the gains of work satisfaction. So, you help the more vulnerable, the very sick, and that feels like a small investment you can make in order to contribute to society. I was very proud of my work at that time.” (participant 96116)

From outside the hospital participants also felt gratitude and respect in the form of banners, messages, fruit baskets and gifts that were sent to the wards. There were two negative cases in this theme: two participants explained that they did not see any role for themselves. They felt that, in a time of life and death, AHPs should exercise restraint and not place themselves in the foreground.

> “In matters of life and death, what should an occupational therapist come around for? It was like, family became very important, as did social work and spiritual guidance, they are very important. It was very medical and then, well, within the entire ICU, at that level of care, there is nothing for you to do as an occupational therapist.” (participant 48827)

Other AHPs helped turning patients on the ICUs, because there were many patients being mechanically ventilated in prone position. They were staffed to assist nurses, where they are used to being in the lead. Some participants explain that this was weird and confusing. It was not clear what was expected from them, and what role they should take. They questioned: What role should I take? AHPs also had questions regarding their identity, for example: Am I a general healthcare professional or a specialized AHP? As a general healthcare professional, they particularly wanted to support nurses and physicians to reduce their workload. As an AHP, they critically considered what care ought to be provided.

> “On the ICU you usually take the lead as a physiotherapist and the nurse has a supportive role and now, we had to, well, search for a new division of roles in which the nurse actually wanted to take the lead. I noticed, especially in that first week, that the nurses were really taken aback and it took a bit of getting used to, like oh, the tube might come off and no, you can’t touch anything. […] And as you are a physiotherapist at heart, you will not simply turn the patient over but you also look at the overall mobility to get a bit of a feeling. Because you don’t know that [patient] population either.” (participant 40447)

Sometimes, based on their skill and knowledge, AHPs saw room for improvement in the situation, however they were not sure whether feedback was appreciated in this situation. They were aware of the workload among the nurses and did not want to be an additional burden. Some participants ensured not to delegate work to nurses on the floor. On the other hand, participants had difficulty not seeing the situation through their allied health lenses. The new disease also raised some professional curiosity. They wanted to add their knowledge and skills where they thought this could be beneficial to the patient. They felt pride in their profession and sought recognition for it. Several participants pointed out that they felt room to take on the expert role during the COVID-19 crisis. They took this period as an opportunity to show other disciplines what they were capable of. For instance, the dieticians helped determine the food demand of ICU patients, in order to relieve ICU physicians from this task. It felt like a victory when this was established and appreciated. In several different ways participants felt room to take up roles that they were normally not able to. Some were national experts and taught webinars. Others experienced that they could finally have substantive talks with physicians. The hierarchy seemed to vanish. Moreover, in the crisis period a lot of things could be organized within short periods of time. These things together made the period energizing, exciting and instructive.

> “The theme leader nutritionist discussed with a privacy officer whether we could also view the patients’ files without referral. And because it was to the benefit of the patient’s treatment, I think we actually received approval for it that same day. When usually you have to get approval from 3 committees, so to speak, it was now arranged within a day. So that is something that changed with Covid-19, now things could be arranged more quickly.” (participant 83109)
>
> “All of a sudden, a lot was approved that would usually take months or years. Protocols were suddenly added on the portal within a week.” (participant 98542)

It was curious to see all the possibilities and opportunities in this period. This made it possible to get things done that had long been desired. This felt refreshing and gave an energy boost. Participants wished they could keep aspects of this new vibe. After the crisis period participants felt recognition and goodwill on the wards. However, some participants pointed out that the hierarchy and bureaucratic procedures started to come back, which felt frustrating.

> “Now we are back to normal and I actually had that feeling already after 3 weeks at the ICU when I thought, ah, okay, we have returned to our regular hierarchy.” (participant 40447)

## Discussion

This qualitative study, using interpretative phenomenological analysis to explore the lived experiences of AHPs working in a hospital during the COVID-19 pandemic revealed four themes: ‘a disease with great impact’, ‘personal health and safety’, ‘staying human in chaotic times’ and ‘solidarity and changing professional roles’. AHPs felt welcomed and appreciated at the COVID-19 wards and ICUs, and were proud that they were able to fulfill their roles. The themes and accompanying ethical issues reflect a wide range of situations that were morally complex and led to moral distress.

Participating AHPs indicated that, during the first wave of the COVID-19 crisis in the Netherlands, the virus had a great impact on the in-hospital working environment due to the massive downscaling of regular care, infection prevention measures and unknown risks to AHPs’ personal health. Normal structures, frameworks, protocols, agreements, roles and certainties did not meet the crisis requirements. This was frightening for the AHPs and came with a need for structure and leadership. New work structures and guidelines were developed, generating an overkill of information. The AHPs had trouble to distinguish main from side issues and to distil what information was actually relevant for their work. At the same time, participants experienced a certain freedom, which made room for authentic motives, connection and solidarity. In the chaotic situations AHPs were urged to rely on their intuition and started acting accordingly. AHPs felt a calling, experienced a sense of being needed and felt which parts of their work were particularly meaningful such as the therapy for patients and support of nurses. These aspects were pointed to as being beautiful, inspiring and giving lots of energy. Organizational changes could be arranged within short periods of time, changes that were impossible to achieve under normal circumstances. AHPs were keen on sharing their expertise and were professionally interested to treat this new patient group as well as possible. Despite the tough conditions and isolation precautions, they sought ways to stay human in line with their fundamental attitude to care for people and to engage in human interactions. AHPs hoped that these meaningful changes from the crisis period would remain. However, they recognized that when the crisis waned, previous hierarchy and bureaucratic procedures seemed to reappear.

In our findings, we recognize two ways of working. Working according to system rules, guidelines or structures and working in a life-world with intuition, intrinsic motivation, trying to do the right thing. These two ways of working show similarities to the two worlds described in Harry Kunneman’s concept of normative professionalism.[19] The system world is the “hard” side of the organization, such as policy forming, planning and control cycles, rules, guidelines and management systems. The system intends that, when professionals comply with the rules, the organization will function properly. The life-world is the world of communication, the world where professionals make decisions based on their own knowledge and experience. The life-world applies to the experiences where professionals determine what to do through communication with each other and with the patient. Both worlds are always in place, however, they require a balance and ideally complement each other. The findings in this study indicate that, to enable healthy deployment of AHPs, the system should not be overemphasized and leave room for the life-world of professionals. This finding seems to touch a fundamental aspect of normative professionalism and might not only be relevant in crisis situations.

The concept of normative professionalism is about “good work”, it is strongly based on Joan Tronto’s ethic of Care. She describes care as: “a species activity that includes everything that we do to maintain, continue, and repair our ‘world’ so that we can live in it as well as possible”.[20] In 1993 Tronto describes four moral qualities as elements of good care: ‘attentiveness’, ‘responsibility’, ‘competence’ and ‘responsiveness’.[20] In the AHPs’ experiences we recognize that all four elements came under pressure during the COVID-19 crisis. For example, AHPs could not fully live up to the quality of ‘competence’, because they had little knowledge about the disease, and they had to fulfill other roles than before. The ethical issues faced by AHPs also reflect straining of these moral qualities. In 2013 Tronto adds a fifth moral quality to her vision: solidarity.[21] This solidarity was strongly evoked during the COVID-19 crisis, and made most AHPs experience a sense of being needed. These findings are in line with earlier studies reporting that healthcare professionals overall felt a strong sense of duty to work during a pandemic influenza,[22] and during the COVID-19 crisis in China.[23]

In the participant experiences in this study several types of moral distress as described by Morley et al. can be recognized.[24] Moral dilemma distress was present when AHPs experienced the dilemma that they wanted to help nurses and physicians, however, they did not know whether it was safe to work in the hospital, and they did not want to put their loved ones at risk. Moral values such as professional loyalty and personal health were at stake, because participating in health care would be loyal though it could lead to viral spreading to loved ones. Moral uncertainty distress was present in the lack of knowledge of the disease, expressed in the values competence, beneficence and non-maleficence, relating to AHPs insecurity about how to treat patients in the best way. They did not dare to trust their clinical experience. Moral constraint distress occurred in relation to values such as professionalism, responsibility, and duty. For instance when AHPs were not able to treat patients due to isolation restrictions and shortages in PPE material, or when professionals could not support patients in situations of loneliness or fear due to visitation restrictions. Moral constraint distress also occurred when patients had to be seen using e-consulting, restricting their clinical view and not allowing them to physically examine patients, which felt like inferior treatment. However, patients might have experienced e-consulting differently, as several studies have indicated that patient experiences and outcomes from telemedicine are comparable to standard practice.[25-27]

Moral distress is a natural response to morally difficult encounters in the provision of patient care.[24] Ethical conflicts and moral distress will always exist in healthcare, especially in high-intensity settings with ethical decision-making.[28] To avoid long-lasting negative impact of moral distress, efforts should be made to mitigate its effects. Until now, only few evidence-based interventions have been studied with limited effectiveness.[29] Yet, some promising practices can be suggested. [24,28,30] The recently developed SUPPORT model enables organizations to simultaneously develop ethical skills and facilitate team-based dialogue while also creating policies shaped by standards of healthy work environments.[28] Based on this model, ethical issues should be recognized and acknowledged. Ethical dialogue should be normalized, and safety should be established for discussions among team members. The organization should encourage debriefing and create conditions to engage in ethical reflection. The importance of ethical reflection is also stressed in Kunneman’s concept of normative professionalism.[19] It enables professionals to create room and reflect on “slow questions”.[31] Slow questions concern life questions about relations, health, loss, violence or longings, issues that cannot be solved by quick technological solutions. The ethical issues faced by AHPs in the COVID-19 crisis were morally complex, requiring a pragmatic tradeoff of values in the search for good care, and therefore, needing recognition and acknowledgement.

Limitations of this study include the lack of data triangulation. The use of other data collection methods, for instance participatory observations, could have enriched the data. However, due to the limited availability of PPE, and the risk of virus transmission this was not an option. Risk of viral spreading was also the reason that interviews had to be performed through video calling. Video calling might have limited the richness of the interviews because of less rapport between interviewer and participant and less non-verbal communication. However, recent studies suggest that in-person interviews are only marginally superior to video calls.[32,33] For this study only the perspective of AHP employees was sought, while the perspectives and interests of managers of organizations, or politicians countrywide, might be opposing. All study participants were AHPs employed in Dutch hospitals. The generalizability of findings might be limited since AHPs in other countries might have had different roles and might have been working under different circumstances. Furthermore, the severity of the COVID-19 crisis was different between countries and might therefore not be comparable. However, the curve of hospitalizations was similar to other countries, at least in Western Europe.

In conclusion, during the COVID-19 crisis, AHPs faced a wide range of situations that were morally complex and led to moral distress, requiring a pragmatic tradeoff of values in the search for good care, and therefore, needing recognition and acknowledgement. To diminish long-lasting negative impact of the COVID-19 pandemic and moral distress, employers should empathize with experiences of allied health professionals and create conditions for ethical reflection. Our data show that allied health professionals value professional autonomy. Creating room for professional autonomy makes them feel needed, connected and energized. However, the needs of allied health professionals may conflict with organizational rules and structures.

## Data Availability

Data are digitally secured at the Radboudumc and available upon reasonable request.

## Abbreviations

COVID-19: Coronavirus Disease 19
AHP: allied health professional
ICU: intensive care unit
PPE: personal protection equipment

## Acknowledgements

We would like to thank all interview participants for their generous participation.

## Data availability statement

Data are digitally secured at the Radboudumc and available upon reasonable request.

## Author contributions

The study was designed by RvO and NK. Data was collected by RvO, EK and NK. Data was analyzed by RvO and NK and checked by AO. The manuscript was written by RvO, AO and NK. MvdB, HK, HV, MG and PvdW contributed to data analysis in the consensus meetings and provided feedback on preliminary manuscript drafts. All authors gave approval for submission of the final version.

## References

1. Dong E, Du H, Gardner L. An interactive web-based dashboard to track COVID-19 in real time. The Lancet infectious diseases. 2020;20(5):533–534.

2. NICE. https://www.stichting-nice.nl/covid-19-op-de-zkh.jsp.

3. Thunström L, Newbold SC, Finnoff D, et al. The benefits and costs of using social distancing to flatten the curve for COVID-19. Journal of Benefit-Cost Analysis. 2020:1–27.

4. Ovadia K, Gazit I, Silner D, et al. Better late than never: a re-examination of ethical dilemmas in coping with severe acute respiratory syndrome. Journal of Hospital Infection. 2005;61(1):75–79.

5. Kole J, de Ruyter D. Werkzame idealen: Ethische reflecties op professionaliteit. Uitgeverij Van Gorcum; 2007.

6. Keulartz J, Schermer M, Korthals M, et al. Ethics in technological culture: a programmatic proposal for a pragmatist approach. Science, Technology, & Human Values. 2004;29(1):3–29.

7. Tigard DW. Rethinking moral distress: conceptual demands for a troubling phenomenon affecting health care professionals. Medicine, Health Care and Philosophy. 2018;21(4):479–488.

8. Campbell SM, Ulrich CM, Grady C. A broader understanding of moral distress. Moral Distress in the Health Professions: Springer; 2018. p. 59–77.

9. Oh Y, Gastmans C. Moral distress experienced by nurses: a quantitative literature review. Nursing ethics. 2015;22(1):15–31.

10. Whitehead PB, Herbertson RK, Hamric AB, et al. Moral distress among healthcare professionals: Report of an institution-wide survey. Journal of Nursing Scholarship. 2015;47(2):117–125.

11. Husserl E, Kersten F. Ideas Pertaining to a Pure Phenomenology and to a Phenomenological Philosophy.: General Introduction to Pure Phenomenology. 1985.

12. Smith JA, Shinebourne P. Interpretative phenomenological analysis. American Psychological Association; 2012.

13. O’Brien BC, Harris IB, Beckman TJ, et al. Standards for reporting qualitative research: a synthesis of recommendations. Academic Medicine. 2014;89(9):1245–1251.

14. Bevan MT. A method of phenomenological interviewing. Qualitative health research. 2014;24(1):136–144.

15. Van Manen M. Phenomenology of practice: Meaning-giving methods in phenomenological research and writing. Routledge; 2016.

16. Association WM. Declaration of Helsinki. Ethical principles for medical research involving human subjects. Jahrbuch Für Wissenschaft Und Ethik. 2009;14(1):233–238.

17. Guideline IHT. Guideline for good clinical practice. J Postgrad Med. 2001;47(3):199–203.

18. van Oorsouw R, Oerlemans A, Klooster E, et al. A sense of being needed: an interpretative phenomenological analysis of hospital-based allied health professionals′ experiences during the COVID-19 pandemic. medRxiv. 2020.

19. Kunneman H. Good work: The ethics of craftsmanship. Uitgeverij SWP; 2013.

20. Tronto JC. Moral boundaries: A political argument for an ethic of care. Psychology Press; 1993.

21. Tronto JC. Caring democracy: Markets, equality, and justice. NYU Press; 2013.

22. Ives J, Greenfield S, Parry JM, et al. Healthcare workers’ attitudes to working during pandemic influenza: a qualitative study. BMC public health. 2009;9(1):1–13.

23. Liu Q, Luo D, Haase JE, et al. The experiences of health-care providers during the COVID-19 crisis in China: a qualitative study. The Lancet Global Health. 2020.

24. Morley G, Sese D, Rajendram P, et al. Addressing caregiver moral distress during the COVID- 19 pandemic. Cleveland Clinic journal of medicine. 2020.

25. Cottrell MA, Galea OA, O’Leary SP, et al. Real-time telerehabilitation for the treatment of musculoskeletal conditions is effective and comparable to standard practice: a systematic review and meta-analysis. Clinical rehabilitation. 2017;31(5):625–638.

26. Tenforde AS, Hefner JE, Kodish-Wachs JE, et al. Telehealth in physical medicine and rehabilitation: a narrative review. PM&R. 2017;9(5):S51–S58.

27. Howard IM, Kaufman MS. Telehealth applications for outpatients with neuromuscular or musculoskeletal disorders. Muscle & nerve. 2018;58(4):475–485.

28. Pavlish C, Brown-Saltzman K, So L, et al. SUPPORT: an evidence-based model for leaders addressing moral distress.JONA: The Journal of Nursing Administration. 2016;46(6):313–320.

29. Dacar SL, Covell CL, Papathanassoglou E. Addressing moral distress in critical care nurses: a systemized literature review of intervention studies. Connect: The World of Critical Care Nursing. 2019;13(2):71–89.

30. Rushton CH, Schoonover-Shoffner K, Kennedy MS. Executive summary: transforming moral distress into moral resilience in nursing. Journal of Christian Nursing. 2017;34(2):82–86.

31. Kunneman H. Voorbij het dikke-ik Amsterdam. Netherlands: BV Uitgeverij SWP. 2005.

32. Irani E. The Use of Videoconferencing for Qualitative Interviewing: Opportunities, Challenges, and Considerations. SAGE Publications Sage CA: Los Angeles, CA; 2019.

33. Krouwel M, Jolly K, Greenfield S. Comparing Skype (video calling) and in-person qualitative interview modes in a study of people with irritable bowel syndrome–an exploratory comparative analysis. BMC medical research methodology. 2019;19(1):219.

